# Real-World Effectiveness of Osteoporosis Treatments on Bone Strength and the Impact of Sequential Anabolic Therapy

**DOI:** 10.64898/2026.10.23.26366354

**Authors:** Koji Ishikawa, Tony M. Keaveny, Xiaodong Li, Yifei Shi, David C. Lee, Soji Tani, Tomoko Towatari, Benjamin D. Elder, Peter G. Passias, Mary Oates

## Abstract

**Summary:** In this real-world biomechanical CT analysis study, romosozumab showed greater increases in vertebral bone strength and density than teriparatide and denosumab in treatment-naïve women with postmenopausal osteoporosis. Romosozumab also increased bone strength after prior teriparatide, whereas twice-weekly teriparatide failed to maintain gains achieved with prior romosozumab.

**Purpose:** Evidence regarding the comparative effectiveness of osteoporosis treatments in improving bone strength in clinical practice remains limited. This real-world study compared the effectiveness of different osteoporosis treatments in women with postmenopausal osteoporosis (PMO), including treatment-naïve patients initiating Romo, TPTD, or DMAb (Study 1) and patients switching from one anabolic therapy to another (Study 2).

**Methods:** Japanese women with PMO aged >55 years were included. In Study 1, 57, 34, and 50 women received Romo, TPTD, or DMAb, respectively, for 12 months. In Study 2, 16 received Romo for 12 months after TPTD (TPTD-Romo) and 29 received twice-weekly TPTD for 12 months after Romo (Romo-TPTD). The primary and secondary endpoints were average percent change from baseline at 12 months in vertebral bone strength and volumetric BMD (vBMD), respectively, assessed using biomechanical CT analysis (BCT).

**Results:** At baseline, overall bone strength and vBMD were comparable across groups in both studies. In Study 1, average percent change (95% CI) in vertebral strength was highest with Romo (39.5% [34.2, 44.7]), followed by TPTD (24.4% [18.8, 30.0]), and DMAb (10.9% [7.6, 14.2]). In Study 2, vertebral strength increased by 16.1% (10.6, 21.6) with TPTD-Romo, and decreased by 7.7% ( 10.7, 4.6) with Romo-TPTD despite subsequent TPTD treatment. Similar trends were observed for vertebral vBMD.

**Conclusion:** In this real-world BCT study, Romo showed greater increases in both vertebral bone strength and density versus TPTD and DMAb in treatment-naïve women with PMO. Among prior TPTD, Romo further increased bone strength and density; however, twice-weekly TPTD failed to maintain the gains following prior Romo.

## INTRODUCTION

The primary goal of osteoporosis treatment is to improve bone strength and reduce fracture risk; appropriate initial treatment selection is essential for optimizing patient outcomes. Different classes of antiresorptive agents, denosumab (DMAb) and bisphosphonates, are widely used in clinical practice to increase bone mineral density (BMD) and reduce fracture risk, although their mechanism of action differ [1–5]. In contrast, anabolic agents such as romosozumab (Romo) have emerged as more potent therapeutic options, as they increase BMD more rapidly and reduce the risk of fractures compared with antiresorptive agents [6–11]. This efficacy is attributed to Romo’s dual mechanism of action which simultaneously stimulates bone formation and inhibits bone resorption [7, 12]. A meta-analysis of 10 randomized controlled trials involving 15,476 patients demonstrated that Romo produced significantly greater increases in lumbar spine BMD compared with placebo, DMAb, teriparatide (TPTD), or alendronate [13]. However, changes in lumbar spine BMD by DXA may not reflect treatment-specific changes in vertebral strength [14]. Thus, evidence regarding the relative effectiveness of Romo compared with other treatments in increasing bone strength in real-world clinical practice remains limited [15].

Although anabolic therapies demonstrate strong efficacy, given their limited treatment duration, sequential treatment plays a critical role in achieving sustained fracture prevention. Previous studies have shown that sequential treatment with alendronate or DMAb following Romo prevents BMD decline and reduce fracture risk [6, 16]. However, these antiresorptive agents generally do not induce further rapid increases in BMD, which may be necessary in patients who remain at very high risk of fracture [6, 16]. In such cases, switching to another anabolic agent may offer additional benefits by promoting greater and more rapid improvements in BMD and bone strength. In clinical practice, however, sequential anabolic therapy is uncommon, and its effects on bone strength remain poorly understood. While several studies have reported that Romo increases BMD following prior TPTD treatment, the use of TPTD after prior Romo has shown limited efficacy in increasing BMD [17, 18]. Furthermore, the impact of sequential anabolic therapy on bone strength has not been investigated.

Although BMD correlates with bone strength, it does not fully capture changes in bone microarchitecture or distinguish treatment including effects on cortical and trabecular bone compartments [19]. Recently, the U.S. Food and Drug Administration qualified total hip BMD as a validated surrogate endpoint for assessing therapeutic efficacy in clinical trials of osteoporosis treatments [20]. However, accurate assessment of vertebral fracture risk based on lumbar spine BMD remains challenging [21, 22]. Lumbar spine BMD measured by dual-energy X-ray absorptiometry (DXA) can be substantially affected by degenerative changes, including osteophytes, facet arthropathy, endplate sclerosis, and aortic calcification, limiting the accuracy of two-dimensional DXA in assessing true vertebral bone strength [23–25]. Opportunistic CT based Hounsfield unit measurements can provide complementary information, but they are derived from selected axial slices, primarily reflect trabecular rather than cortical bone, and are susceptible to technical variation, which may limit sensitivity to some treatment related changes [15]. In contrast, bone strength estimated by finite element analysis using biomechanical computed tomography (BCT) provides a functional, three-dimensional assessment that directly reflects the mechanical properties of bone and serves as a more clinically relevant predictor of fracture risk [10, 26].

## PURPOSE

In this study, we evaluated the real-world effectiveness of osteoporosis treatments on vertebral bone strength using BCT–based finite element analysis. Our primary objective was to compare the effects of Romo, TPTD, and DMAb, commonly used injectable osteoporosis therapies in patients at high risk of fracture. In addition, given the limited duration of anabolic therapy and the clinical importance of treatment sequencing, we investigated the impact of 12 months of subsequent anabolic treatment strategies, particularly switching between Romo and TPTD in women who received prior anabolic treatment, to determine whether such treatment sequences provide additional benefits in improving vertebral bone strength.

## METHODS

### Study design and population

This retrospective observational study in Japan included women with PMO (age > 55 years) at high risk of fracture. Patients were included if they completed 12 months of treatment with Romo (210 mg once monthly), TPTD (20 μg once daily or 28.2 μg twice weekly), or DMAb (60 mg once every 6 months) and had CT scans available both within 60 days before treatment initiation and at 12 months after treatment initiation. Patients were excluded if they had (1) secondary osteoporosis, (2) poorly controlled thyroid disease, (3) active malignancy, or (4) conditions affecting the musculoskeletal system, such as Parkinson’s disease. Based on prior treatment history, patients were classified into two studies, “Study 1 (treatment naïve)” and “Study 2 (anabolic switch)”.

In Study 1, the treatment-naïve cohort included patients who had not received osteoporosis treatment within 3 years before treatment initiation. In Study 2, the anabolic-switch cohort included patients who had received TPTD or Romo for at least 12 months and then switched to the other anabolic agent (Fig 1).

**Fig 1.**
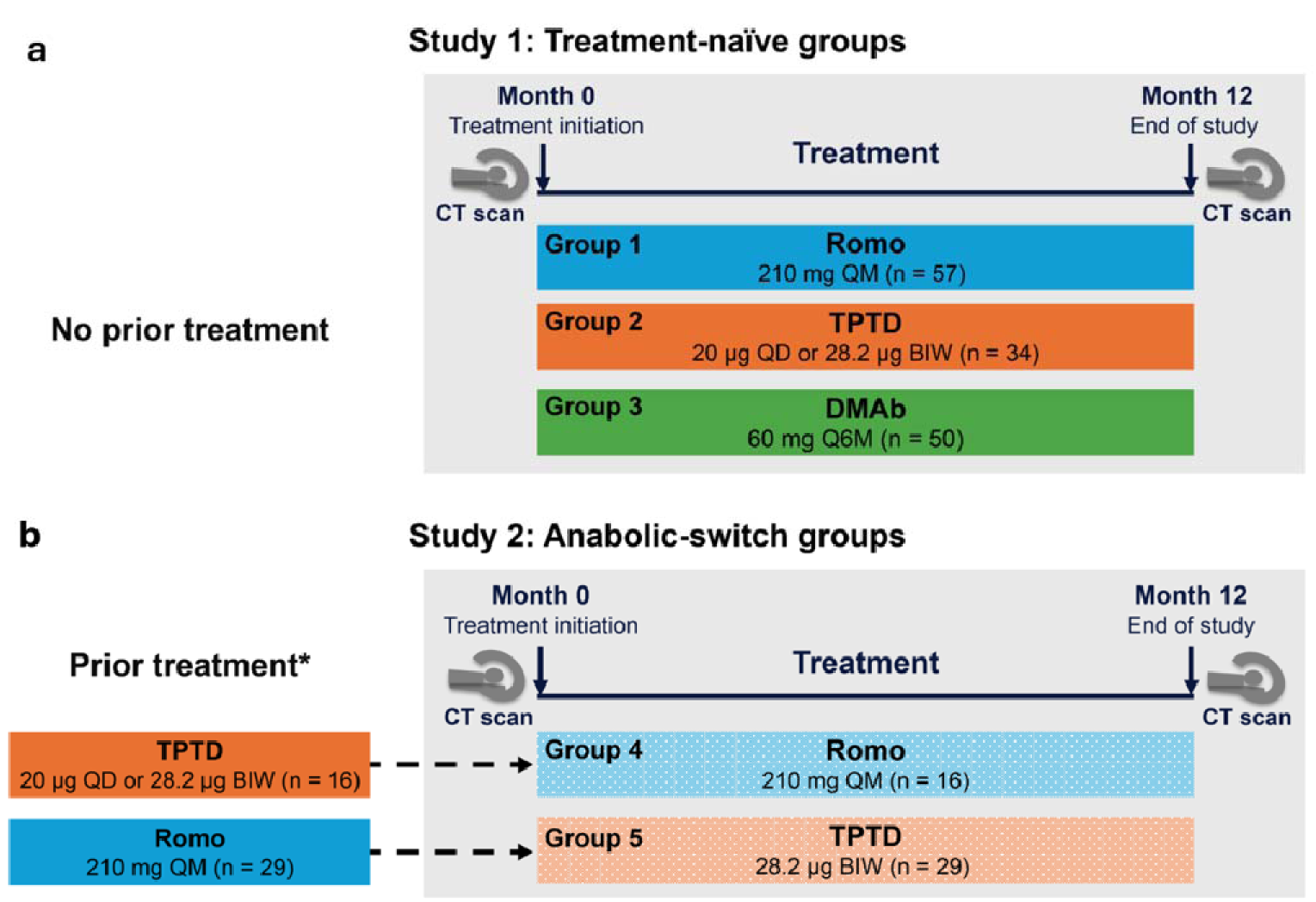
Study design. BIW, twice a week; DMAb, denosumab; QD, once daily; Q6M, once every 6 months; QM, once every month; Romo, romosozumab; TPTD, teriparatide. (a) Study design for study 1 treatment-naïve groups where patients received no osteoporosis treatment for 3 years before treatment initiation. (b) Study desgin for study 2 anabolic-switch groups. *Mean duration of prior treatment: TPTD = 20 months; Romo = 12 months. In both the studies, CT scans were taken within 2 months before treatment initiation and at 12 (±1) months.

The study protocol was reviewed and approved by an institutional review board (IRB) (IRB number: #24102206-4). This study involved secondary analysis of data from the Fujigoko cohort, which was designed to investigate the effects of osteoporosis treatment. All participants provided written informed consent. Only de-identified data were transferred and analyzed for this study. Therefore, additional informed consent was not required according to local governmental ethical guidelines for epidemiological research. The study was conducted in accordance with relevant ethical guidelines for research involving human participants.

### BCT analyses

CT scan data were acquired using a GE Revolution EVO ES scanner and Toshiba Aquilion scanner under predefined scanning conditions consistent with our previous studies: 120 kVp, slice thickness ≤ 2mm, with a standard reconstruction kernel [9, 24, 25, 27]. In total, scans for 186 patients (both from Studies 1 and 2) were included, with spine and/or femur coverage at baseline and month 12. To ensure consistency in longitudinal measurements, BMD was primarily calibrated using a standard external phantom (Mindways, Austin, TX, USA) beneath the patient. For a subset of 50 patients where protocol variables such as scanner vendor or table height differed, a phantomless calibration approach was utilized to maintain calibration stability [28]. By referencing internal tissues adjacent to the bone rather than to an external material, phantomless calibration effectively mitigates the effects of scan field variations, ensuring consistent BMD measurements across the cohort. BCT was applied to these CT data to measure finite-element analysis derived bone strength and volumetric BMD (vBMD), each at the hip and spine. BCT analysis was performed using VirtuOst^®^ software (version 2.9, O.N. Diagnostics, Berkeley, CA, USA) as previously described [10, 26]. Briefly, following image segmentation, calibration, registration, and resampling, the target bones from the scans were converted into finite element models by transforming each voxel into 1.0-mm-cube–shaped elements, resulting in models with approximately 20,000–50,000 elements in the vertebral models and 80,000–160,000 in the femur models. Using these models, vertebral strength and vBMD were assessed, for the whole bone and the cortical and trabecular compartments. Femoral strength and vBMD were also assessed. All analyses were performed blinded to treatment status.

### Study endpoints

The primary endpoint of the study was to estimate the least squares mean (LSM) percent change from baseline in vertebral strength at 12 months. Secondary endpoints were LSM percent change from baseline at 12 months for the following parameters: vertebral trabecular and cortical (outer 2 mm of bone) compartment strength; vertebral integral vBMD including trabecular and cortical compartment vBMD; and proximal femoral strength and vBMD.

### Statistical Analyses

Statistical analyses were descriptive, with treatment effects summarized using point estimates and 95% confidence intervals (CIs). Baseline demographics and patient characteristics were summarized descriptively by treatment group. Categorical data were summarized by the number and percentage of patients in each treatment group with non-missing observations. Continuous data were summarized as means and standard deviation (SD). The LSM percent change from baseline and difference between the groups for the study endpoints were compared using a generalized linear model (GLM). The GLM model was adjusted for the baseline value of respective endpoint, patient’s age, and patient’s fracture history before treatment initiation. In the analysis of the study endpoints, precision around point estimates was described using 95% CIs.

## RESULTS

### Study 1 (treatment-naïve)

#### Baseline characteristics

At treatment initiation, mean (SD) age of patient was not different across the Study 1 treatment-naïve groups (Romo, 73.8 [7.9] years; TPTD, 74.1 [7.6] years; and DMAb, 75.1 [8.6] years) (Table 1). Approximately 80% of patients in the Romo and TPTD groups and 50% in the DMAb group had prior fracture history (Table 1). At baseline, mean (SD) bone strength at spine was 3680 (840) N, 3510 (1040) N, and 3740 (1180) N among the Romo, TPTD, and DMAb groups, respectively (Table 1). Similarly, baseline cortical or trabecular vertebral compartment strength were numerically similar across these treatment-naïve groups. Average vertebral integral vBMD and cortical or trabecular vertebral compartment vBMD were comparable across groups. Bone strength and vBMD at the proximal femur were numerically similar between the Romo and TPTD groups and were numerically lower in the DMAb group (Table 1).

**Table 1.** Baseline characteristics of treatment-naïve groups.

| **Characteristics** | **Romo**<br>**(N = 57)** | **TPTD**<br>**(N = 34)** | **DMAb**<br>**(N = 50)** |
| --- | --- | --- | --- |
| Age (years) | 73.8 (7.9) | 74.1 (7.6) | 75.1 (8.6) |
| Fracture history, n (%) | 45 (79) | 27 (79) | 25 (50) |
| Vertebral strength (N) | 3680 (840) | 3510 (1040) | 3740 (1180) |
| Vertebral cortical compartment strength (N) | 2060 (550) | 1900 (490) | 2020 (560) |
| Vertebral trabecular compartment strength (N) | 1630 (550) | 1610 (620) | 1720 (690) |
| Vertebral integral vBMD (mg/cm<sup>3</sup>) | 132 (20) | 127 (23) | 129 (27) |
| Vertebral cortical compartment vBMD (mg/cm<sup>3</sup>) | 184 (29) | 174 (24) | 174 (29) |
| Vertebral trabecular compartment vBMD (mg/cm<sup>3</sup>) | 106 (18) | 102 (23) | 105 (26) |
| Proximal femoral strength (N) | 2440 (600) | 2420 (570) | 2060 (600) |
| Proximal femoral vBMD (mg/cm<sup>3</sup>) | 201 (43) | 196 (43) | 179 (44) |

DMAb, denosumab; N, newtons, Romo, romosozumab; TPTD, teriparatide; vBMD, volumetric bone mineral density.
Continuous measures are expressed in mean (SD).

#### Treatment effects at the spine

After 12 months of treatment, the LSM percent change (95% CI) in vertebral strength from baseline was highest at 39.5% (34.2, 44.7) with Romo, followed by 24.4% (18.8, 30.0) with TPTD and 10.9% (7.6, 14.2) with DMAb (Fig 2, Table S1). The LSM difference (95% CI) in vertebral bone strength was 14.4% (8.3, 20.6) between the Romo and TPTD groups and 26.5% (20.7, 32.2) between the Romo and DMAb groups. Similarly, increases in trabecular and cortical compartment vertebral strength were highest with Romo, followed by TPTD and DMAb. The LSM difference (95% CI) in trabecular compartment strength was 11.3% (2.5, 20.1) for Romo versus TPTD and 26.9% (18.6, 35.2) for Romo versus DMAb. Similarly, the LSM difference (95% CI) in cortical compartment strength was 18.8% (13.2, 24.4) for Romo versus TPTD and 27.4% (22.1, 32.7) for Romo versus DMAb.

**Fig 2.**
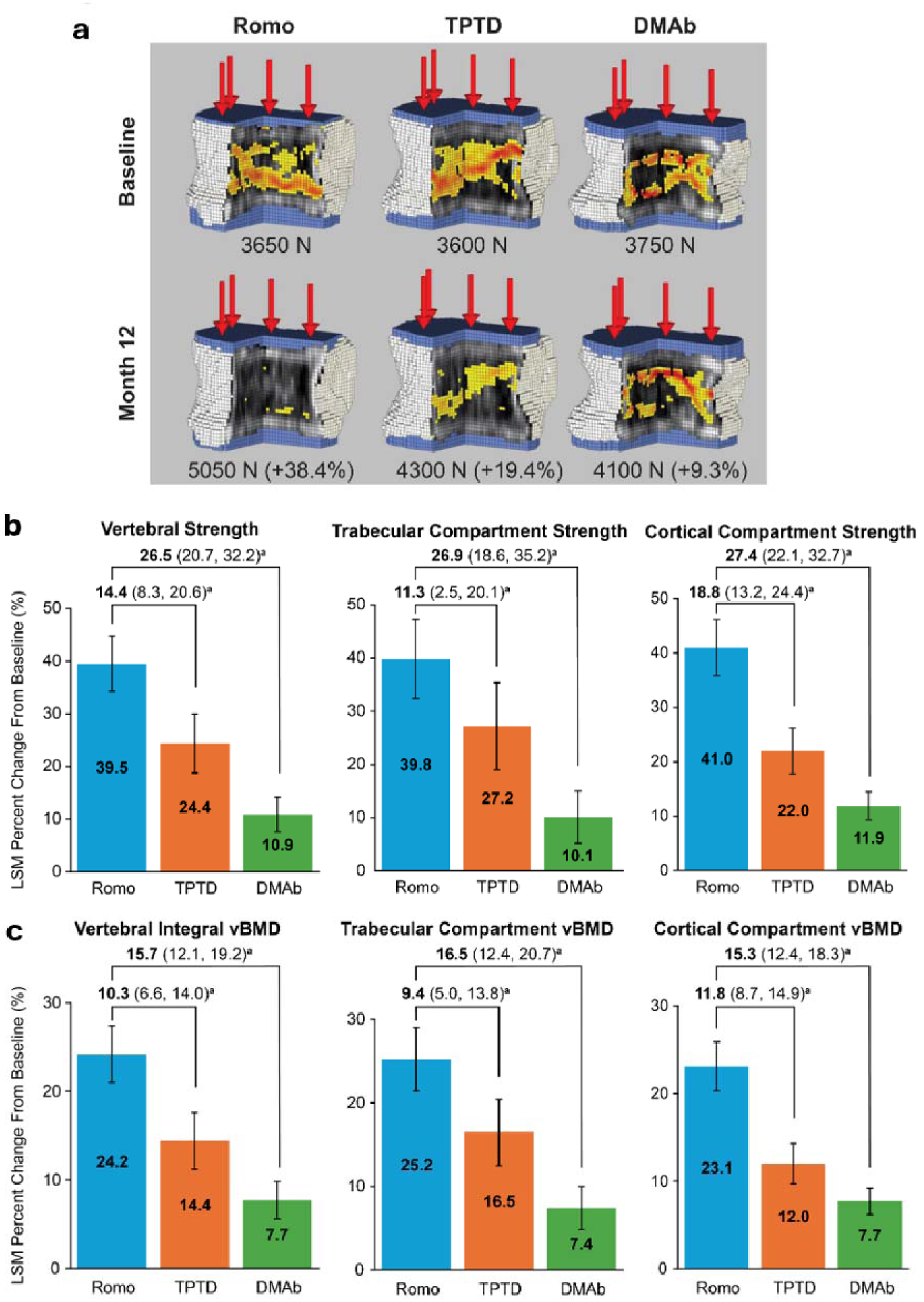
Changes in vertebral bone strength and vBMD after 12 months of treatment initiation among treatment-naïve patients. BCT, biomechanical computed tomography; Cl, confidence interval; DMAb, denosumab; LSM, least squares mean; Romo, romosozumab; TPTD, teriparatide; vBMD, volumetric bone mineral density. (a) Representative images of the spine were selected based on similar values of baseline vertebral strength. Yellow-red color map = regions of failed tissue after virtual loading. LSM percent change (95% CI) from baseline at month 12 in (b) vertebral strength, including trabecular and cortical compartment strength, and (c) vertebral integral vBMD, including trabecular and cortical compartment vBMD. Vertebral endpoints were based on evaluable BCT scans: Romo, *n* = 50; TPTD, *n* = 33; DMAb, *n* = 49. ^a^LSM difference estimate between groups (95% CI).

The LSM percent change (95% CI) in vertebral integral vBMD from baseline was 24.2% (21.0, 27.4) with Romo, 14.4% (11.2, 17.6) with TPTD, and 7.7% (5.6, 9.8) with DMAb (Fig 2, Table S1). The LSM difference (95% CI) was 10.3% (6.6, 14.0) between the Romo and TPTD groups and 15.7% (12.1, 19.2) between Romo and DMAb groups. Similarly, increase in trabecular and cortical compartment vertebral vBMD was highest with Romo followed by TPTD and DMAb (Fig 2). The LSM difference (95% CI) for trabecular and cortical compartment vBMD was 9.4% (5.0, 13.8) and 11.8% (8.7, 14.9), respectively, between the Romo and TPTD groups and 16.5% (12.4, 20.7) and 15.3% (12.4, 18.3), respectively, between the Romo and DMAb groups.

#### Treatment effects at the hip

Overall, the increase in bone strength and vBMD was less at the proximal femur as compared with the spine. LSM percent change (95% CI) in proximal femoral strength from baseline was 7.4% (5.2, 9.6) with Romo, 4.4% (2.1, 6.8) with TPTD, and 4.7% (2.9, 6.5) with DMAb (Fig S1, Table S1). The LSM difference (95% CI) was 2.7% (0.0, 5.4) between the Romo and TPTD groups and 3.3% (0.5, 6.1) between the Romo and DMAb groups. The LSM percent change (95% CI) in proximal femoral vBMD from baseline was 5.4% (4.0, 6.9) with Romo, 3.4% (1.9, 5.0) with TPTD, and 3.6% (2.5, 4.7) with DMAb (Fig S1). The LSM difference (95% CI) was 2.2% (0.4, 4.0) between the Romo and TPTD groups and 2.3% (0.6, 4.1) between the Romo and DMAb groups.

### Study 2 (anabolic-switch)

#### Baseline characteristics

At treatment initiation, mean (SD) age of patients in Study 2 (anabolic-switch) was 74.4 (7.7) years and 73.9 (8.0) years in the TPTD-Romo and Romo-TPTD groups, respectively (Table 2). Approximately, 80% of women in the TPTD-Romo group and 60% in the Romo-TPTD group had prior fracture history (Table 2). Average (SD) spine bone strength at baseline was comparable between the two anabolic-switch groups (TPTD-Romo, 5070 [980] N; Romo-TPTD, 5050 [1340] N; Table 2), but both switching groups had overall higher bone strength than treatment-naïve groups in Study 1, likely reflecting the effects of prior treatment. Baseline cortical (TPTD-Romo, 2650 [540] N; Romo-TPTD, 2770 [740] N) or trabecular (TPTD-Romo, 2420 [580] N; Romo-TPTD, 2280 [720] N) compartment vertebral strengths were similar between the switching groups. Average (SD) vertebral integral vBMD was comparable between the two switching groups (TPTD-Romo, 156 [20] mg/cm^3^; Romo-TPTD, 165 [28] mg/cm^3^) and numerically higher than that reported in the study 1 (treatment-naïve) groups. Similarly, baseline cortical or trabecular vertebral compartment vBMD were numerically similar between the switching groups and higher than those in the treatment-naïve groups. At baseline, bone strength at the proximal femur was numerically higher in the TPTD-Romo group (mean [SD], 2810 [500] N) than in the Romo-TPTD group (mean [SD], 2480 [420] N), while vBMD was similar between the two switching groups (Table 2).

**Table 2.** Baseline characteristics of “anabolic-switch” groups.

| Status of Prior Treatment | Prior TPTD | Prior Romo |
| --- | --- | --- |
| Characteristics | Romo<br>(N = 16) | TPTD<br>(N = 29) |
| Age (years) | 74.4 (7.7) | 73.9 (8.0) |
| Fracture history, n (%) | 13 (81) | 17 (59) |
| Vertebral strength (N) | 5070 (980) | 5050 (1340) |
| Vertebral cortical compartment strength (N) | 2650 (540) | 2770 (740) |
| Vertebral trabecular compartment strength (N) | 2420 (580) | 2280 (720) |
| Vertebral integral vBMD (mg/cm<sup>3</sup>) | 156 (20) | 165 (28) |
| Vertebral cortical compartment vBMD (mg/cm<sup>3</sup>) | 207 (20) | 225 (33) |
| Vertebral trabecular compartment vBMD (mg/cm<sup>3</sup>) | 131 (20) | 134 (27) |
| Proximal femoral strength (N) | 2810 (500) | 2480 (420) |
| Proximal femoral vBMD (mg/cm<sup>3</sup>) | 220 (28) | 204 (34) |

Continuous measures are expressed in mean (SD).

#### Treatment effects at the spine

Among the anabolic-switch groups, after 12 months of Romo or TPTD therapy in women with prior TPTD or Romo treatment, the LSM percent change (95% CI) in vertebral strength increased by 16.1% (10.6,21.6) from baseline in the TPTD-Romo group whereas it decreased by 7.7% ( 10.7, 4.6) in the Romo-TPTD group (Fig 3, Table S2).

**Fig 3.**
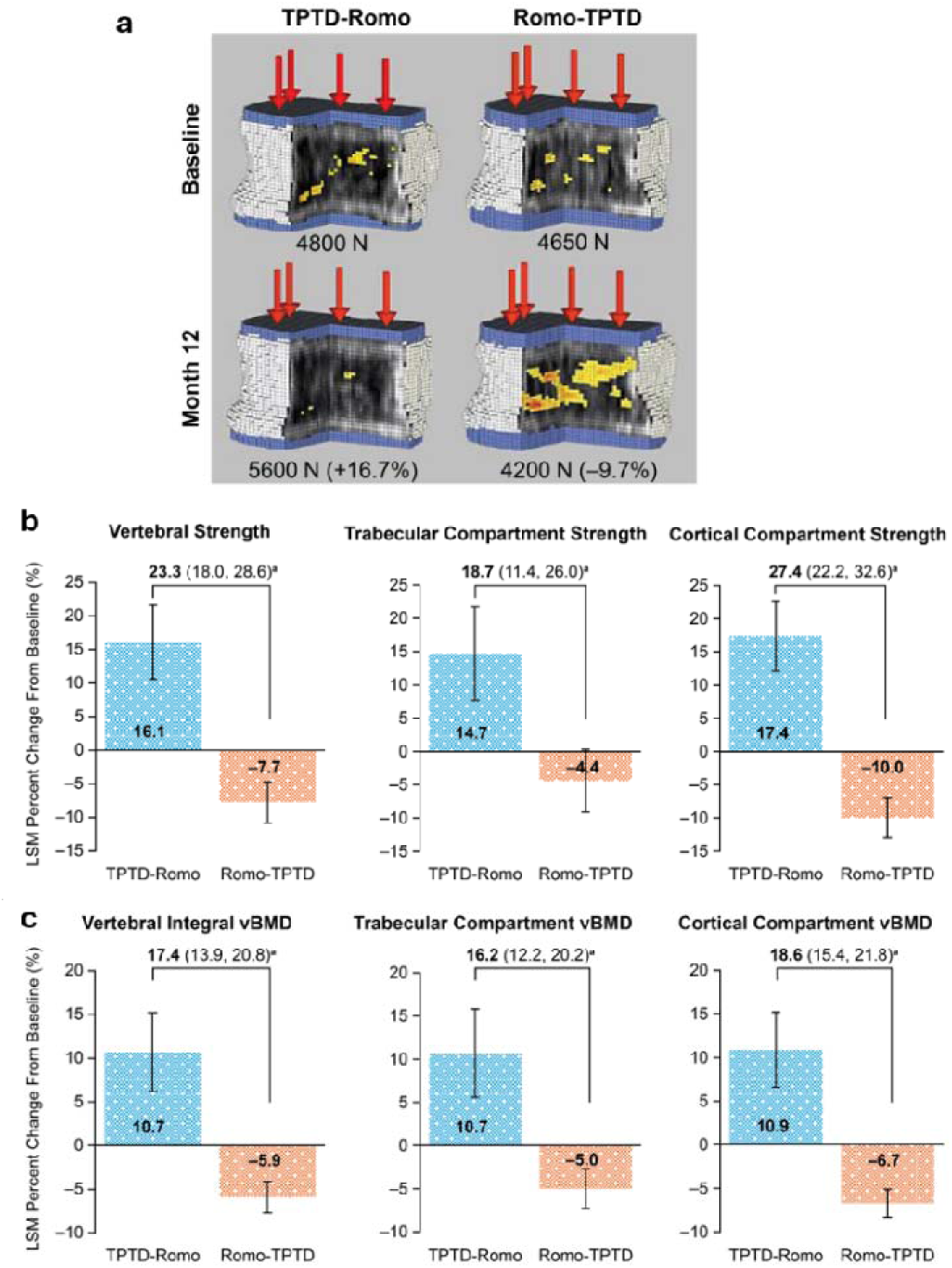
Changes in vertebral bone strength and vBMD after 12 months of treatment initiation among patients in the “anabolic-switch” groups. BCT, biomechanical computed tomography; Cl, confidence interval; DMAb, denosumab; LSM, least squares mean; Romo, romosozumab; TPTD, teriparatide; vBMD, volumetric bone mineral density. (a) Representative images of the spine were selected based on similar values of baseline vertebral strength. Yellow-red color map = regions of failed tissue after virtual loading. LSM percent change (95% CI) from baseline at month 12 in (b) vertebral strength, including trabecular and cortical compartment strength, and (c) vertebral integral vBMD, including trabecular and cortical compartment vBMD. Vertebral endpoints were based on evaluable BCT scans, TPTD-Romo, *n* = 15; Romo-TPTD, *n* = 26. ^a^LSM difference estimate between groups (95% CI).

The LSM difference (95% CI) was 23.3% (18.0, 28.6) between the TPTD-Romo and Romo-TPTD groups (Fig 3). Trabecular and cortical vertebral compartment strength increased in the TPTD-Romo group while decreased in the Romo-TPTD group. The LSM percent change (95% CI) in vertebral integral vBMD from baseline was 10.7% (6.2, 15.3) in the TPTD-Romo group and –5.9% (–7.7, –4.1) in the Romo-TPTD group, and the LSM difference (95% CI) was 17.4% (13.9, 20.8) (Fig 3). The LSM difference (95% CI) was 16.2% (12.2, 20.2) for trabecular vertebral compartment vBMD and 18.6% (15.4, 21.8) for cortical vertebral compartment vBMD (Fig 3).

#### Treatment effects at the hip

The LSM percent change (95% CI) in proximal femoral strength from baseline was 1.6% (–0.5, 3.7) in the TPTD-Romo group and –0.9% (–2.6, 0.8) in the Romo-TPTD group (Fig S2), with the LSM difference (95% CI) of 2.1% (–0.6, 4.8) (Fig S2, Table S2). The LSM percent change (95% CI) from baseline in vBMD was 1.3% (–0.2, 2.8) in the TPTD-Romo group and –1.2% (–1.9, –0.4) in the Romo-TPTD group, with the LSM difference (95% CI) of 2.9% (1.5, 4.2) (Fig S2).

## DISCUSSION

This is the first real-world CT based study assessing bone strength in Japanese women with PMO in two clinical settings: Study 1, which included treatment-naïve patients treated with Romo, TPTD, or DMAb; and Study 2, which included patients who switched between Romo and TPTD. In treatment-naïve patients (Study 1), Romo was the most effective in increasing overall bone strength and vBMD at both the spine and hip, followed by TPTD and DMAb. In the sequential anabolic therapy group (Study 2), Romo treatment after prior TPTD increased vertebral bone strength and integral vBMD beyond the gains induced by prior TPTD; however, TPTD treatment after prior Romo failed to maintain the gains induced by prior Romo. BCT analysis further demonstrated consistent treatment-related changes across both trabecular and cortical compartments, providing compartment-specific insight beyond whole-bone assessment.

As direct measurement of bone strength is not feasible in clinical practice, CT-based biomechanical analyses, including BCT, have recently been used to estimate strength-related parameters for the evaluation of treatment response and fracture risk [10, 14, 29]. A review of several clinical studies showed that this approach has greater statistical sensitivity than DXA for early detection of bone loss, that CT-derived bone strength is more strongly associated with vertebral fractures than DXA-derived BMD, and that it may be more effective for monitoring treatment response in individual patients [26]. Thus, biomechanical CT assessment could offer clinically meaningful insight into treatment effects.

Our real-world data showed that Romo produced the greatest increase in bone strength among treatment-naïve patients, consistent with prior trial-based evidence. A phase 2 substudy demonstrated greater gains in spine and femur strength with Romo than with TPTD after 12 months of treatment [10]; similarly, the ARCH substudy showed greater improvements in bone strength with Romo than with alendronate [14]. Importantly, our study extends on these findings by assessing treatment effects in a real-world setting, incorporating DMAb into the comparison, analyzing a larger biomechanical CT-based cohort, and examining anabolic-switch groups.

Although DMAb has previously been shown to increase bone strength compared with placebo, [29] our results suggested that its effect on bone strength after 1 year of treatment is lower than that of Romo in clinical practice at both the spine and hip.

After treatment with a bone-forming agent, sequential therapy with bisphosphonates or DMAb is recommended for patients with osteoporosis to maintain or further improve BMD [1]. However, sequential therapy with bisphosphonates does not always produce significant additional BMD gains [30]. As an alternative approach, bone-forming agents have been used sequentially in some patients. Prior studies have shown that this strategy can increase BMD, although the magnitude of the effect is strongly influenced by prior treatment [17, 18, 31–33].

For example, a recent study showed that the percentage increase in areal BMD assessed by DXA at the spine and hip was higher in patients treated with Romo after prior TPTD than in those treated with TPTD after prior Romo therapy (lumbar spine, 10.8% vs 0.0%; total hip, 4.4% vs −1.3%) [17]. Extending these observations, our study suggested that TPTD may be a less favorable sequential option after Romo, given that bone strength gains achieved with prior Romo were not maintained. Interestingly, a prior study suggested that this difference may be partly explained by baseline differences at the time of treatment transition, with lower BMD at the switch from TPTD to Romo allowing greater subsequent gains, whereas little additional BMD increase was observed with TPTD after Romo [17]. However, in our study, bone strength and vBMD at baseline were comparable between the TPTD-Romo and Romo-TPTD groups, yet greater improvements in both parameters were observed in the TPTD-Romo group. These findings suggested that the differences between the two treatment sequences cannot be explained solely by baseline bone strength and vBMD. One possible explanation is the difference in bone-turnover dynamics between the two sequences. Previous studies evaluating changes in bone turnover markers suggest that switching from TPTD to Romo maintains bone formation while reducing bone resorption, potentially resulting in a more favorable anabolic window [17, 31, 32]. In contrast, this window may be less pronounced after switching from Romo to TPTD because bone resorption transiently increases after the switch. These differences may help explain why TPTD did not maintain bone strength after prior Romo treatment. Further studies, including bone histomorphology, may help support this hypothesis.

The main strength of our study is that it is the first real-world study assessing treatment effects on bone strength in clinical practice in a relatively large cohort. In contrast, previous evidence on bone strength have been derived from well-controlled clinical trials such as STRUCTURE, ARCH, and FREEDOM, which have specific predefined patient eligibility criteria (such as fracture history, prior treatment) and high treatment adherence among patient populations, and thus, findings from these clinical trials are less generalizable [10, 11, 14, 29]. Because our study reflected routine clinical practice rather than trial conditions, its findings may be more directly applicable to everyday clinical decision making.

Our study has several limitations. First, its retrospective observational design precludes causal inference, and residual confounding may remain. However, CT-based bone strength analyses were performed by investigators blinded to treatment groups. Between-group comparisons were conducted using multivariable general linear models adjusted for relevant baseline characteristics, with the results presented as adjusted least-squares mean percent changes and 95% CIs. Second, the smaller anabolic-switch cohort limited the precision of the estimates, although an apparent difference was observed between the groups. Despite these limitations, to our knowledge, this is one of the largest real-world cohort studies using CT-based bone strength analysis.

## CONCLUSION

In this real-world BCT study, Romo, TPTD, or DMAb improved bone strength and vBMD in women with PMO. However, the magnitude of improvement varied, with Romo showing greater increase in bone strength and vBMD than TPTD or DMAb. In women who received prior therapy, Romo use after prior TPTD increased bone strength and vBMD; however, twice weekly TPTD failed to maintain the gains in vertebral strength and vBMD induced with prior Romo. Overall, our findings suggest that initial treatment with Romo is the most effective in increasing bone strength and vBMD.

## AUTHOR CONTRIBUTIONS

Koji Ishikawa: Conceptualization, Investigation, Methodology, Data curation, Formal analysis, Writing – original draft, Writing – review & editing. Tony M. Keaveny: Writing – review & editing. Xiaodong Li: Conceptualization, Formal Analysis, Writing – original draft, Writing – review & editing. Yifei Shi: Formal analysis, Writing – review & editing. David C. Lee: Formal analysis, Writing – review & editing. Soji Tani: Data curation, Formal analysis, Writing – review & editing. Tomoko Towatari: Data curation, Formal analysis, Writing – review & editing.

Benjamin D. Elder: Writing – review & editing. Peter G. Passias: Writing – review & editing. Mary Oates: Writing – review & editing

## Data availability

Qualified researchers may request data from Amgen clinical studies. Complete details are available at the following: https://www.ext.amgen.com/science/clinical-trials/clinical-data-transparency-practices/clinical-trial-data-sharing-request/

## Funding

This study was funded by Amgen Inc. and UCB.

## Conflicts of interest disclosure

**KI** has served as a consultant to and received research grants from Amgen Inc. **TMK** has served as a consultant to Amgen Inc., Bone Health Technologies, and Angitia and holds equity in O.N. Diagnostics. **YS and MO** are employees and stockholders of Amgen Inc. **XL** was an employee and stockholder of Amgen Inc. at the time of the study and is currently employed by Angitia Biopharmaceuticals, Westlake Village, California, USA. **DCL** is an employee of O.N. Diagnostics. **ST and TT** declare no conflicts of interest. **BDE** serves on the editorial board of JNS Spine; has served as a consultant to SI-BONE, DePuy Synthes, Amgen Inc., Medtronic, and Iota Biosciences; receives royalties from SI-BONE, Elsevier, and Biomendex; has received clinical trial funding from Stryker, SI-BONE, and Medimaps; serves on the medical advisory board of Injectsense; and holds stock in Injectsense. **PGP** serves on the editorial board of *JNS Spine* and as Deputy Editor of Spine; has received research grants from Globus, Medtronic, and Cerapedics; and receives royalties from Royal Biologics).

## ACKNOWLEDGMENTS

This study was sponsored by Amgen Inc., and UCB Pharma. We thank Ayano Oyamada, Kei Gonsho and Miho Mochizuki for their assistance in collecting clinical data. We thank Lisa Humphries, PhD, Varsha Jain, PhD, and Sneha Sinha, PhD, for medical writing and editorial support (Amgen Inc.)

## Notes

### Author Declarations

The Institutional Review Board of Yamanashi Red Cross Hospital gave ethical approval for this work (IRB approval no. 24102206-4).

