## Supplemental materials for "Real-World Effectiveness of Osteoporosis Treatments on Bone Strength and the Impact of Sequential Anabolic Therapy"

### Supplementary Material

**Table S1.** Observed bone strength and vBMD after 12 months of treatment initiation and percent change from baseline among patients in the "treatment-naive" groups

#### A) Romo versus TPTD

| Characteristics | **Romo (*N* = 57)** | | **TPTD (*N* = 34)** | | **Romo vs TPTD** |
|---|---|---|---|---|---|
| | **Observed value at month 12** | **LSM percent change (95% CI) at month 12** | **Observed value at month 12** | **LSM percent change (95% CI) at month 12** | **LSM percent difference (95% CI)** |
| Vertebral strength (N) | 5020 (1160) | 39.5 (34.2, 44.7) | 4280 (1110) | 24.4 (18.8, 30.0) | 14.4 (8.3, 20.6) |
| Vertebral cortical compartment strength (N) | 2830 (670) | 41.0 (35.9, 46.1) | 2300 (490) | 22.0 (17.8, 26.2) | 18.8 (13.2, 24.4) |
| Vertebral trabecular compartment strength (N) | 2190 (730) | 39.8 (32.3, 47.2) | 1990 (710) | 27.2 (19.0, 35.4) | 11.3 (2.5, 20.1) |
| Vertebral integral vBMD (mg/cm$^3$) | 162 (25) | 24.2 (21.0, 27.4) | 143 (25) | 14.4 (11.2, 17.6) | 10.3 (6.6, 14.0) |
| Vertebral cortical compartment vBMD (mg/cm$^3$) | 225 (31) | 23.1 (20.3, 25.8) | 195 (24) | 12.0 (9.7, 14.4) | 11.8 (8.7, 14.9) |
| Vertebral trabecular compartment vBMD (mg/cm$^3$) | 131 (24) | 25.2 (21.4, 29.0) | 117 (26) | 16.5 (12.6, 20.5) | 9.4 (5.0, 13.8) |
| Proximal femoral strength (N) | 2610 (600) | 7.4 (5.2, 9.6) | 2520 (580) | 4.4 (2.1, 6.8) | 2.7 (0.0, 5.4) |
| Proximal femoral vBMD (mg/cm$^3$) | 212 (43) | 5.4 (4.0, 6.9) | 202 (44) | 3.4 (1.9, 5.0) | 2.2 (0.4, 4.0) |

#### B) Romo versus DMAb

| Characteristics | Romo ($N = 57$) | | DMAb ($N = 50$) | | Romo vs DMAb |
|---|---|---|---|---|---|
| | **Observed value at month 12** | **LSM percent change (95% CI) at month 12** | **Observed value at month 12** | **LSM percent change (95% CI) at month 12** | **LSM percent difference (95% CI)** |
| Vertebral strength (N) | 5020 (1160) | 39.5 (34.2, 44.7) | 4150 (1470) | 10.9 (7.6, 14.2) | 26.5 (20.7, 32.2) |
| Vertebral cortical compartment strength (N) | 2830 (670) | 41.0 (35.9, 46.1) | 2260 (700) | 11.9 (9.4, 14.5) | 27.4 (22.1, 32.7) |
| Vertebral trabecular compartment strength (N) | 2190 (730) | 39.8 (32.4, 47.2) | 1890 (840) | 10.0 (5.1, 15.0) | 26.9 (18.6, 35.2) |
| Vertebral integral vBMD (mg/cm$^3$) | 162 (25) | 24.2 (21.0, 27.4) | 138 (30) | 7.7 (5.6, 9.8) | 15.7 (12.1, 19.2) |
| Vertebral cortical compartment vBMD (mg/cm$^3$) | 225 (31) | 23.1 (20.3, 25.8) | 187 (32) | 7.7 (6.2, 9.3) | 15.3 (12.4, 18.3) |
| Vertebral trabecular compartment vBMD (mg/cm$^3$) | 131 (24) | 25.2 (21.4, 29.0) | 113 (29) | 7.4 (4.8, 10.0) | 16.5 (12.4, 20.7) |
| Proximal femoral strength (N) | 2610 (600) | 7.4 (5.2, 9.6) | 2150 (600) | 4.7 (2.9, 6.5) | 3.3 (0.5, 6.1) |
| Proximal femoral vBMD (mg/cm$^3$) | 212 (43) | 5.4 (4.0, 6.9) | 184 (43) | 3.6 (2.5, 4.7) | 2.3 (0.6, 4.1) |

N, newtons; Romo, romosozumab; TPTD, teriparatide; vBMD, volumetric bone mineral density.

Continuous measures are expressed in mean (SD). Vertebral endpoints were based on evaluable BCT scans: Romo, $n$ = 50; TPTD, $n$ = 33; DMAb, $n$ = 49. Proximal femoral endpoints were based on evaluable BCT scans: Romo, $n$ = 56; TPTD, $n$ = 34; DMAb, $n$ = 50.

**Table S2.** Observed bone strength and vBMD after 12 months of treatment initiation and percent change from baseline among patients in the "anabolic-switch" groups

| **Characteristics** | **TPTD-Romo (*N* = 16)** | | **Romo-TPTD (*N* = 29)** | | **TPTD-Romo vs Romo-TPTD** |
|---|---|---|---|---|---|
| | **Observed value at month 12** | **LSM percent change (95% CI) at month 12** | **Observed value at month 12** | **LSM percent change (95% CI) at month 12** | **LSM percent difference (95% CI)** |
| Vertebral strength (N) | 5790 (1000) | 16.1 (10.6, 21.6) | 4690 (1470) | −7.7 (−10.7, −4.6) | 23.3 (18.0, 28.6) |
| Vertebral cortical compartment strength (N) | 3070 (530) | 17.4 (12.1, 22.6) | 2510 (790) | −10.0 (−13.0, −7.1) | 27.4 (22.2, 32.6) |
| Vertebral trabecular compartment strength (N) | 2720 (610) | 14.7 (7.7, 21.7) | 2180 (770) | –4.4 (–9.1, 0.3) | 18.7 (11.4, 26.0) |
| Vertebral integral vBMD (mg/cm$^3$) | 173 (26) | 10.7 (6.2, 15.3) | 155 (32) | –5.9 (–7.7, –4.1) | 17.4 (13.9, 20.8) |
| Vertebral cortical compartment vBMD (mg/cm$^3$) | 229 (25) | 10.9 (6.6, 15.3) | 210 (37) | –6.7 (–8.3, –5.1) | 18.6 (15.4, 21.8) |
| Vertebral trabecular compartment vBMD (mg/cm$^3$) | 144 (25) | 10.7 (5.6, 15.7) | 128 (30) | –5.0 (–7.3, –2.8) | 16.2 (12.2, 20.2) |
| Proximal femoral strength (N) | 2860 (520) | 1.6 (–0.5, 3.7) | 2460 (420) | –0.9 (–2.6, 0.8) | 2.1 (–0.6, 4.8) |
| Proximal femoral vBMD (mg/cm$^3$) | 224 (28) | 1.3 (–0.2, 2.8) | 201 (33) | –1.2 (–1.9, –0.4) | 2.9 (1.5, 4.2) |

N, newtons; Romo, romosozumab; TPTD, teriparatide; vBMD, volumetric bone mineral density.

Continuous measures are expressed in mean (SD). Vertebral endpoints were based on evaluable BCT scans: TPTD-Romo, *n* = 15; Romo-TPTD, *n* = 26. Proximal femoral endpoints were based on evaluable BCT scans: TPTD-Romo, *n* = 16; Romo-TPTD, *n* = 29.

**Fig S1** Changes in proximal femoral bone strength and vBMD after 12 months of treatment initiation among treatment-naïve patients

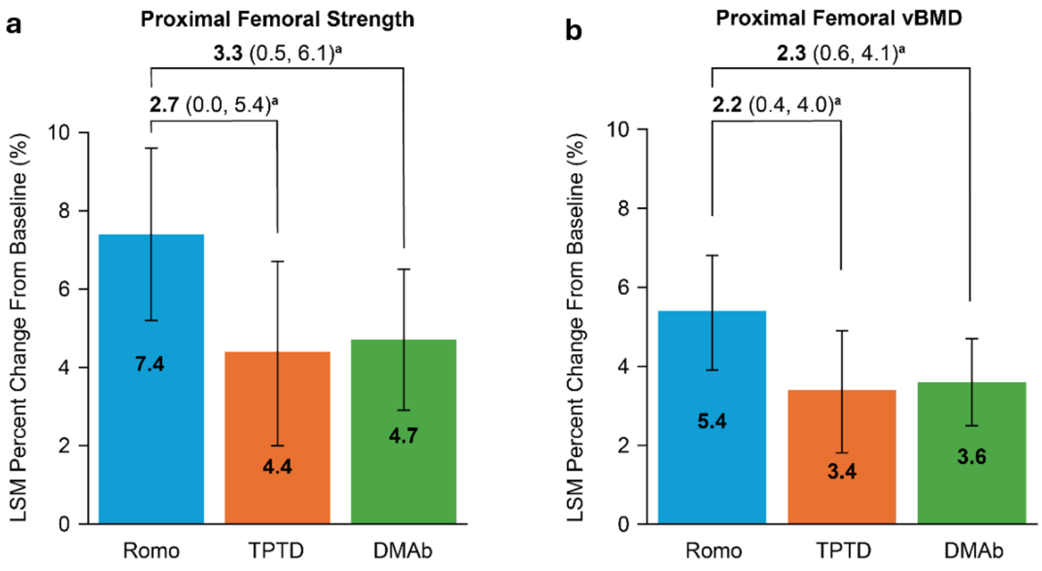


BCT, biomechanical computed tomography; Cl, confidence interval; DMAb, denosumab; LSM, least squares mean; Romo, romosozumab; TPTD, teriparatide; vBMD, volumetric bone mineral density.

LSM percent change (95% CI) from baseline at month 12 in (A) proximal femoral strength, (B) proximal femoral vBMD. Proximal femoral endpoints were based on evaluable BCT scans: Romo, *n* = 56; TPTD, *n* = 34; DMAb, *n* = 50.

[a]LSM difference estimate between groups (95% CI).

**Fig S2** Changes in proximal femoral bone strength and vBMD after 12 months of treatment initiation among patients in the anabolic-switch groups

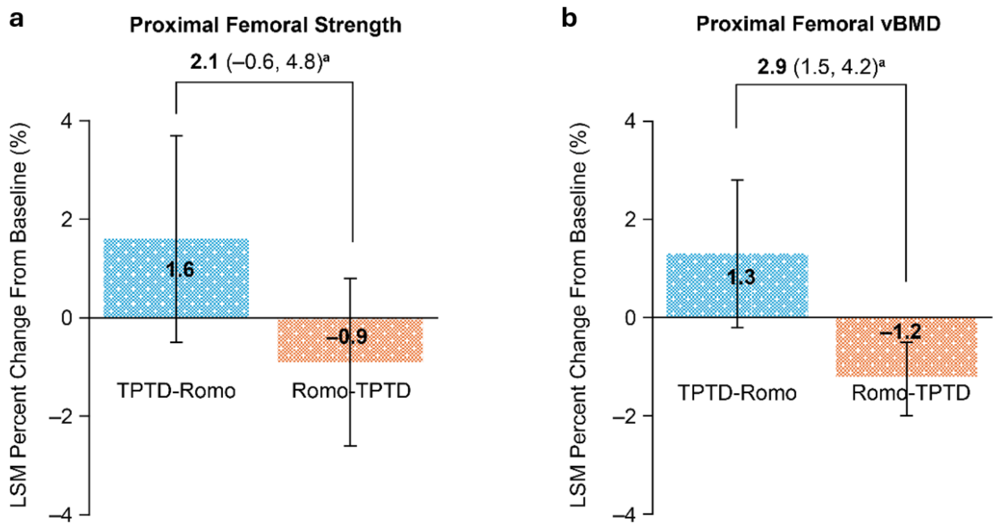


BCT, biomechanical computed tomography; Cl, confidence interval; DMAb, denosumab; LSM, least squares mean; Romo, romosozumab; TPTD, teriparatide; vBMD, volumetric bone mineral density.

LSM percent change (95% CI) from baseline at month 12 in (A) proximal femoral strength, (B) proximal femoral vBMD. Proximal femoral endpoints were based on evaluable BCT scans, TPTD-Romo = 16; Romo-TPTD = 29.

[a]LSM difference estimate between groups (95% CI).

**Fig S3** Baseline characteristics and changes in vertebral and proximal femoral strength and vBMD after 12 months Romo after a prior DMAb treatment

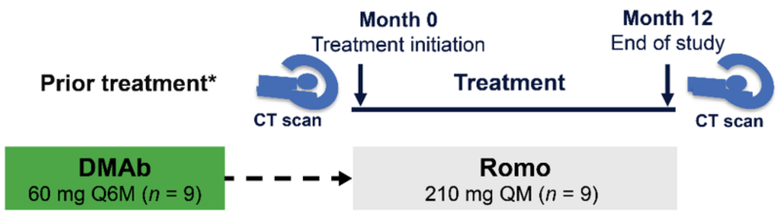


b

| | Romo after DMAb (*N* = 9) |
|---|---|
| Age (years) | 78.6 (6.5) |
| Fracture history, *n* (%) | 8 (89) |
| Vertebral strength (N) | 4360 (1410) |
| Vertebral vBMD (mg/cm$^3$) | 145 (36) |
| Proximal femoral strength (N) | 2620 (620) |
| Proximal femoral vBMD (mg/cm$^3$) | 207 (48) |

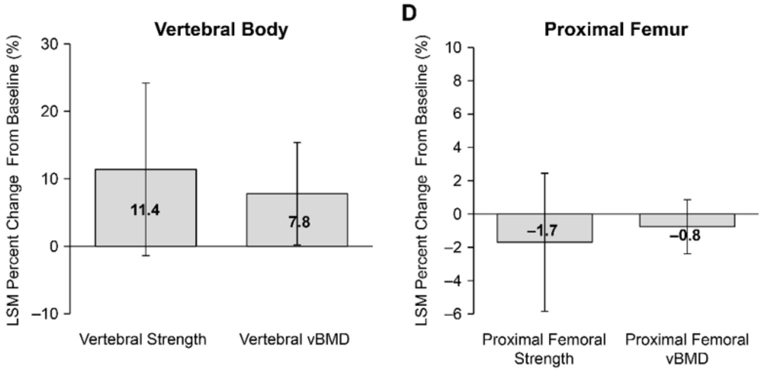


DMAb, denosumab; LSM, least squares mean; PMO, postmenopausal osteoporosis; Q6M, once every 6 months; QM, once every month; Romo, romosozumab; vBMD, volumetric BMD.

(A) Study design for women with PMO who received Romo after a prior treatment with DMAb. *Mean duration of prior treatment: DMAb = 43 months. CT scans were taken within 2 months before treatment initiation and at 12 (±1) months. (B) Baseline characteristics of women with PMO. Continuous measures are expressed in mean (SD).

LSM percent change (95% CI) from baseline at month 12 in vertebral strength and vBMD (C) and proximal femoral strength and vBMD (D).

**Supplementary text**

Among women with postmenopausal osteoporosis (PMO) who received romosozumab (Romo) after prior therapy with denosumab (DMAb; $n$ = 9), 89% had a prior fracture history. At baseline, mean (SD) bone strength at spine was 4360 (1410) N and volumetric bone mineral density (vBMD) was 145 (36) mg/cm$^3$. After 12 months of therapy, the least squares mean (LSM) percent change from baseline was 11.4% in vertebral strength and 7.8% in vBMD. At the hip, the LSM percent change in bone strength was –1.7% and –0.8% in vBMD. Overall, Romo followed by DMAb induced some increase in bone strength and vBMD at the vertebrae but no change at the hip.
